# Low-intensity focused ultrasound stimulation in stroke: An intensity escalation phase I safety and feasibility study

**DOI:** 10.1101/2024.09.12.24313472

**Authors:** Ziping Huang, Charalambos C. Charalambous, Mengyue Chen, Taewon Kim, Estate Sokhadze, Allen Song, Sin-Ho Jung, Shashank Shekhar, Jody Feld, Xiaoning Jiang, Wuwei Feng

**Affiliations:** Department of Neurology, Duke University School of Medicine; Department of Biomedical Engineering, Duke University; Department of Mechanical and Aerospace Engineering, North Carolina State University; Department of Physical Medicine and Rehabilitation, Penn State College of Medicine; Department of Kinesiology, Pennsylvania State University; Duke Brain Imaging and Analysis Center, Duke University School of Medicine; Department of Biostatistics and Bioinformatics, Duke University School of Medicine; Department of Orthopaedic Surgery, Duke University School of Medicine

**Keywords:** low-intensity focused ultrasound stimulation, stroke, motor learning, corticospinal excitability, safety

## Abstract

**BACKGROUND:** Low-intensity focused ultrasound stimulation (LIFUS) has recently emerged as a promising neuromodulation tool for certain neuropsychiatric diseases. However, its safety and feasibility in stroke patients remains unknown. Intensity is a critical safety parameter for LIFUS. We aimed to determine the maximum safe and tolerable intensity of LIFUS in stroke patients, and to explore its effect on upper-extremity motor learning and corticospinal excitability.

**METHODS:** Subjects with first-ever stroke participated in this Phase I study. We adopted the classic 3+3 dose-escalation paradigm to sham/0, 1, 2, 4, 6, and 8 W/cm^2^ spatial-peak pulse-average intensity (I_SPPA_, estimated in-vivo transcranial value; LOW: sham/0, 1 and 2 W/cm^2^, HIGH: 4, 6 and 8 W/cm^2^). Stopping rules (dose limiting toxicities) were pre-defined: ≥2^nd^-degree scalp burn, clinical seizures, ≥20% topical apparent diffusion coefficient change, or participant discontinuation due to any reason. A 12-minute LIFUS was applied over the ipsilesional motor cortex while participants were concurrently practicing three blocks of motor sequence learning (MSL) task using the affected hand. We collected the occurrences of pre-defined adverse events, post-minus-pre improvements in MSL response time, and post-minus-pre differences in corticospinal excitability quantified by motor evoked potentials.

**RESULTS:** I_SPPA_ was escalated to 8 W/cm^2^ with eighteen stroke participants without meeting stopping rules. Compared to the LOW, the HIGH performed significantly better on the MSL (24.7±13.3% vs. 13.2±10.9%, *p*=0.01). Similarly, the HIGH also showed signs of increased corticospinal excitability (32.0±34.3% vs. 12.9±48.0%) but did not reach significance, *p*=0.53.

**CONCLUSIONS:** Our Phase I safety study suggests that a single session of 12-minute LIFUS up to 8 W/cm^2^ I_SPPA_ is safe and feasible in stroke patients. Higher LIFUS intensities can induce greater MSL retention. The next logical step is to conduct a Phase II study to further test the efficacy of LIFUS and monitor its safety profiles in stroke patients.

**REGISTRATION:** URL: https://www.clinicaltrials.gov; Unique identifier: NCT05016531

## INTRODUCTION

Low-intensity focused ultrasound stimulation (LIFUS) is a novel non-invasive neuromodulation tool that uses mechanical acoustic waves. It is capable of stimulating whole-brain depths and millimeter focuses^1^. It also shows promising characteristics of being steerable, targeted, selective and reversible^2,3^. Based on these advantages and the promising data from animal studies and healthy volunteers^1,2,4,5^, we postulated that LIFUS has the potential to be an effective modulatory tool in influencing motor cortex excitabilities which have been implicated in post-stroke motor impairments and central post-stroke pain. However, in order to systematically investigate LIFUS in stroke patients, safety profiles of LIFUS parameters must be determined. Among several ultrasound parameters, the intensity of the acoustic wave, defined as the power carried by sound waves per unit area in the direction perpendicular to that area, influences safety the most. Studies in healthy volunteers used a range of LIFUS intensities without any major safety-related complications^1,4,5^. However, due to the physiological differences between healthy and diseased populations, LIFUS parameters cannot be translated over directly. Therefore, we aimed to conduct a Phase I intensity/dose escalation safety and feasibility study in a cohort of stroke patients. In addition, we explored LIFUS’s effects on motor learning and corticospinal excitability.

## METHODS

### Trial design overview

This Phase I-type dose-escalation study was conducted at Duke University Hospital, Durham, USA. The dose-escalation scheme **(Figure 1A)** is adapted from the well-established 3+3 trial design^6^ that is commonly used to find the maximally tolerable dose for chem-drugs. For LIFUS, the acoustic intensity (spatial-peak pulse-average intensity, I_SPPA_, estimated in-vivo transcranial value) is the dosage parameter for this study.

**Figure 1.**
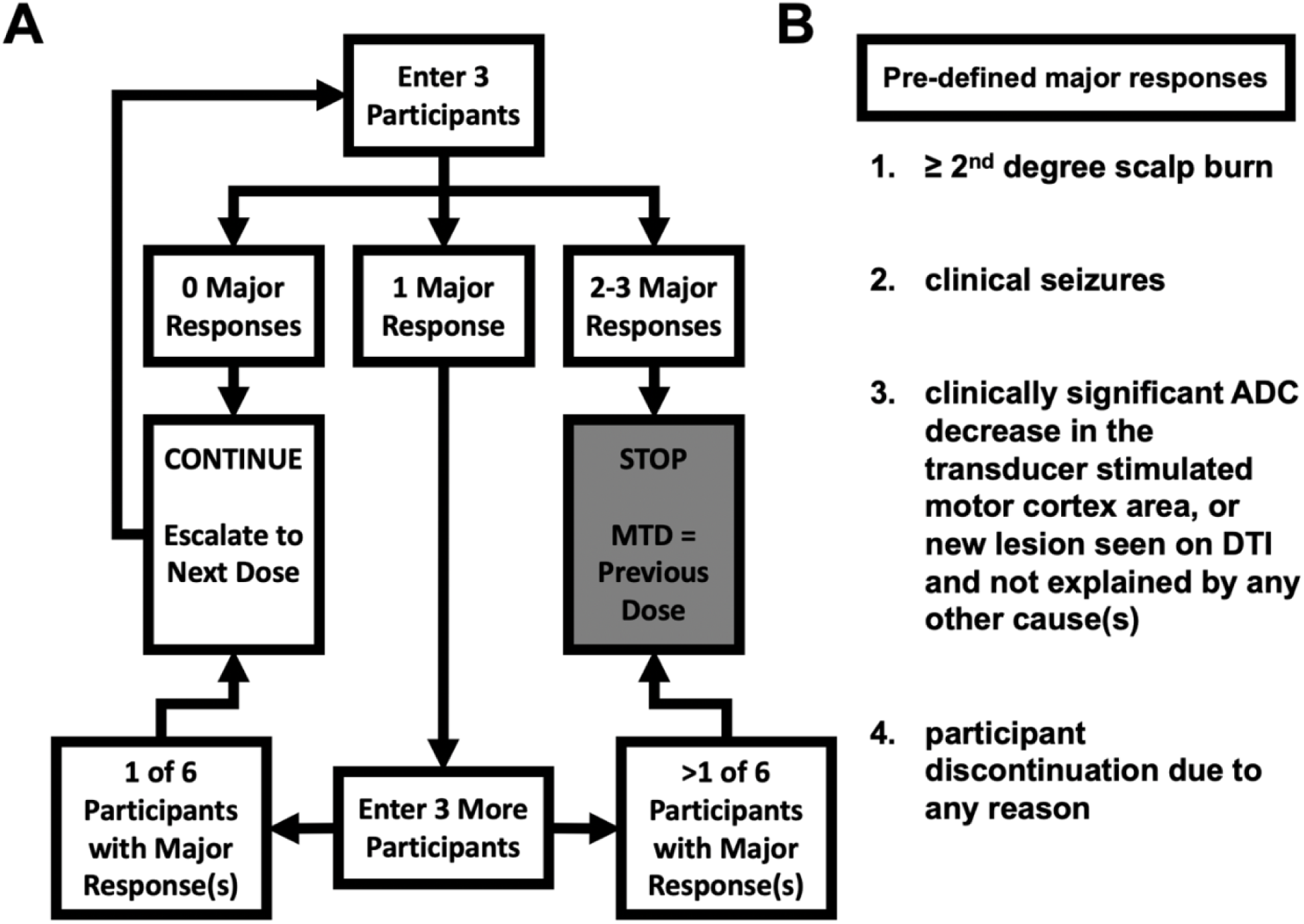
3+3 intensity-escalation design. **A**, Intensity-escalation scheme with incremental intensities of sham/0, 1, 2, 4, 6 and 8 W/cm^2^. **B**, Pre-defined major responses. MTD: maximally tolerable dose; ADC: apparent diffusion coefficient; DTI: diffusion tensor imaging.

Starting with 0 W/cm^2^, our escalation scheme first enrolled three participants, then, (a) if no major response (defined below) occurred in all of these three participants, we would escalate to the next intensity; (b) if two to three participants had a major response, the trial would be terminated; and (c) if major response(s) occurred in only one participant, we would enroll three additional participants at the same intensity: if no major response occurred in these three participants, we would escalate to the next intensity; if any further major response occurred, the trial would be terminated. In either of the two termination scenarios, the one intensity below the terminating dose would be the maximum safe and tolerable intensity.

The intensity escalation regimen was, sham/0 W/cm^2^ >> 1 W/cm^2^ >> 2 W/cm^2^ >> 4 W/cm^2^ >> 6 W/cm^2^ >> 8 W/cm^2^. The pre-defined major responses (i.e., stopping rules; **Figure 1B)** were: 1) ≥ 2^nd^-degree topical scalp burn; 2) clinical seizures; 3) topical apparent diffusion coefficient (ADC) decrease equal or more than the minimal clinically significant 20%^7^ or new lesion(s) not explainable by any other causes seen on diffusion tensor imaging (DTI); and 4) participant asking to be withdrawn for any reason.

### Patient population

Stroke participants were included only if they: 1) were ≥ 21 years old of any race or gender; 2) had a first-ever ischemic or hemorrhagic stroke more than one month ago; 3) had a unilateral limb weakness with a Fugl Meyer-Upper Extremity (FM-UE) score ≤ 62 (out of 66)^8^; and 4) had transcranial magnetic stimulation (TMS)-elicitable motor-evoked potentials (MEPs) on the paretic abductor pollicis brevis (APB). Participants were excluded if they had: 1) any concomitant neurological disorder(s) affecting arm functions; 2) a documented history of severe dementia with or without medications before the stroke; or 3) any contraindication(s) to magnetic resonance imaging (MRI)/TMS/LIFUS^9–11^. The study protocol received approval from the Institutional Review Board at Duke University. All participants gave a written informed consent.

### Experimental procedures

This study consists of one baseline visit and one testing visit **(Figure 2A)**. At the baseline visit, participants gave their written informed consent, followed by determining eligibility and collecting vital signs and clinico-demographic information. A licensed physical therapist assessed upper extremity motor impairment using the FM-UE scale, after which we tested participants for their TMS-MEP status. If eligible, participants underwent anatomical and diffusion MRIs.

**Figure 2.**
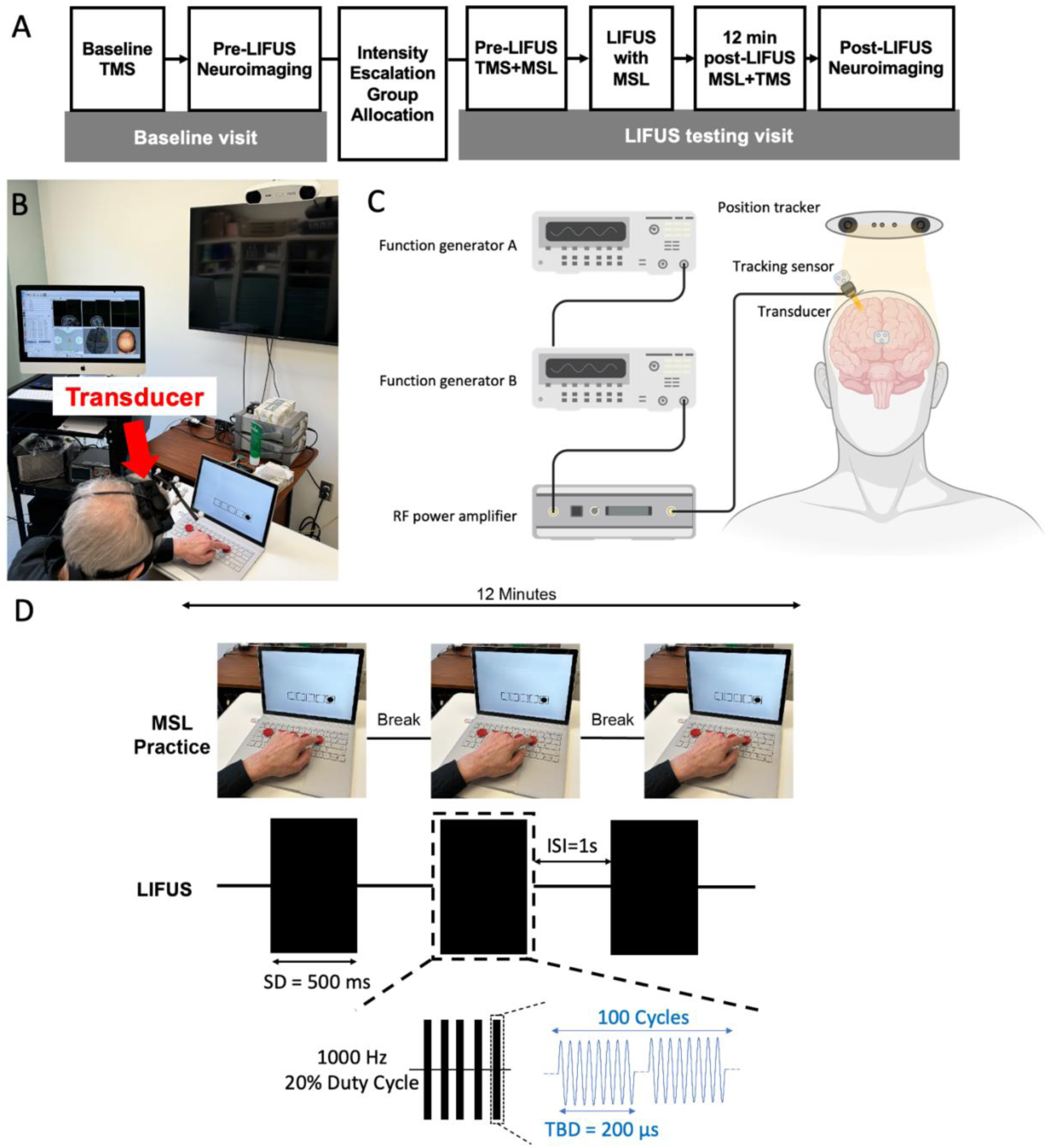
Experimental procedures. **A,** Study visit activities. **B**, Concurrent neuronavigated-LIFUS and MSL setup during the LIFUS administration with an author (and not a study participant) pictured. **C,** System component schematic of the neuronavigated-LIFUS. **D**, Concurrent MSL-LIFUS protocol with an author (and not a study participant) picutred. TMS: transcranial magnetic stimulation; LIFUS: low-intensity focused ultrasound stimulation; MSL: motor sequence learning; RF: radio frequency; SD: sonication duration; ms: milliseconds; ISI: inter-stimulus interval; s: second; Hz: Hertz; TBD: tone burst duration; μs: microseconds.

For the testing visit, participants were instructed to wash their hair prior to their arrival. The testing visit began with a navigation-guided, single-pulse TMS for APB MEPs in a seated position and proceeded with one block of a baseline motor sequence learning (MSL) task. Then, a 12-minute continuous and neuronavigated LIFUS was administered **(Figure 2B**; **Figure 2C)**. Parallelly, participants completed three practice blocks of the same MSL task, interleaved with 1-2 minutes breaks **(Figure 2D)**. After the LIFUS, a study team member first inspected the scalp for any burn/injury. Then participants performed a post-stimulation MSL assessment, followed by a TMS assessment, and lastly another MRI.

### LIFUS protocol

We utilized a custom-made, 500 kHz single-element focused ultrasound transducer with a 30 mm focal depth for stimulations (Blatek Industries, Inc., PA, USA). Two function generators (33210A, Keysight Technologies, Inc., CA, USA) were utilized to generate the desired electrical waveform^12^ **(Figure 2C)**. Specifically, function generator (FG) A delivered 500 pulses at a 1000 Hz pulse repetition frequency (PRF) to trigger FG B, while FG B provided 100 cycles of a 500 kHz sine wave per pulse. Overall, the electrical waveform consists of a 200 μs tone burst duration, a 500 ms sonication duration (SD), and a 1 s inter-stimulus interval **(Figure 2D)**. A radio frequency power amplifier (50A250, Amplifier Research Corporation, PA, USA) amplified the electrical waveform from FG B to drive the ultrasound transducer. Based on our previous transcranial ultrasound wave propagation experiments and simulations^13,14^, we set the measured I_SPPA_ in free water to 40 W/cm^2^, providing the estimated transcranial 8 W/cm^2^ I_SPPA_ considering skull attenuations. Other estimated in-vivo parameters included a 0.69 mechanical index (MI) and a 533 mW/cm^2^ averaged spatial-peak temporal-average intensity (I_SPTA_), all within the Food and Drug Administration (FDA) safety guidance for diagnostic ultrasounds (MI < 1.9, I_SPPA_ < 190 W/cm^2^ and I_SPTA_ < 720 mW/cm^2^)^15^.

For precise stimulations, we mounted the transducer with a Brainsight CT-913 tracking sensor (Rouge Research Inc., Quebec, Canada) for a transducer, participants’ MRI and TMS coil co-registration **(Figure 2B**; **Figure 2C)**. Ultrasound gel (Aquasonic 100, Parker Laboratories, Inc., NJ, USA) was applied between the transducer and the scalp “hot-spot” – the optimal spot where TMS elicited the largest MEP on the contralateral APB. We monitored transducer location online, adjusted its position if necessary, and retrospectively assessed transducer locations offline^16^.

### Neuroimaging protocol

Neuroimaging was collected on a 3-Tesla GE Signa UHP MRI scanner at Duke Brain Imaging and Analysis Center. Three-dimensional T1 magnetization-prepared rapid acquisition gradient-echo (MPRAGE) and T2-weighted fluid attenuated inversion recovery (FLAIR) images (1 mm isotropic spatial resolution) were acquired, along with a DTI sequence (dual spin-echo echo-planar imaging acquisitions, 2 mm isotropic spatial resolution, b factor = 1000 s/mm^2^).

### MSL protocol

We utilized the discrete sequence production task, which has been extensively employed to examine motor sequence learning^17^. We created the MSL task in-house (E-Prime 3.0, Psychology Software Tools, PA, USA), and placed a five-button keyboard (1 cm-diameter round buttons, horizontally equidistant) in front of participants, at a location where participants confirmed comfortable reach with their affected hand. In addition, participants saw five horizontally equidistant, 2 cm x 2 cm boxes on a computer screen, each corresponding to one keyboard button. With a circle appearing in one of the boxes, participants were cued to hit the corresponding button using their stroke-affected hand **(Figure 2D)**. The circle appeared uniquely within a sequence, e.g., box number 4-1-3-5-2. Each participant got assigned a sequence randomly and received this same sequence on their pre-LIFUS assessment (one block), practices during LIFUS (three blocks), and their post-LIFUS assessment (one block) MSL. Each block consisted of twelve repetitions of the participant’s assigned sequence. We delivered all MSL tasks without mentioning “a sequence” in any form, and only asked participants to complete the task as quickly and accurately as possible using just their affected hand.

### TMS protocol

Identical TMS-MEP procedures were performed across participants for a neurophysiological assessment of their corticospinal excitability. Participants sat in a comfortable position with arms and hands at rest. Using Brainsight 2.4.10 (Rouge Research Inc., Quebec, Canada), participants had their physical anatomical landmarks co-registered to the MNI ICBM 152 template^18^ for the baseline screening, and to their T1 neuroimages in the testing session. Electrodes (Kendall^TM^ H124SG, Cardinal Health 200, LLC, IL, USA) were applied to participants’ paretic APB in a belly-tendon montage and were connected to the CED1902 amplifier (Cambridge Electronic Design Limited, Cambridge, UK). We also used their MICRO4 analog-to-digital converter, and their Signal 7.05a (x86) software for data acquisitions. We set the TMS stimulator (BiStim^2^, Magstim Inc, MN, USA) to the BiStim mode, and had their 70 mm figure-of-eight coil oriented for posteroanterior intracranial currents. Single monophasic TMS pulses were bilaterally applied over motor cortical areas to elicit MEPs larger than 50 μV peak-to-peak on the contralateral APB while it was at rest. After the detection of a coil setup that best elicited contralateral APB MEPs, we determined the resting motor threshold (RMT) and testing motor threshold (TMT) with this coil setup (the “hot-spot”) using the adaptive parameter estimation by sequential testing method^19^ (MTAT 2.1). RMT and TMT were defined as the percentage of maximum stimulator output (MSO) required to attain 50 μV and 1 mV MEP, respectively. When a 50 μV MEP was attainable but a 1 mV was not, we used 100% MSO. Then, twenty single pulses spaced over 10 seconds apart^20^ were applied over the “hot-spot” using either the TMT or 100% MSO when the TMT could not be determined. TMS assessments were collected both before and after the LIFUS. The post-stimulation TMS started 12 minutes after LIFUS, for a transducer removal, safety assessments and TMS preparations.

### Data Analyses

#### ADC calculation

To assess topical ADC changes, we corrected DTI images for movements and eddy currents^21^, fitted voxel-wise diffusion tensors^22^, and took the voxel-wise mean diffusivity (MD) to obtain ADC maps. Region of interest (ROI) for ADC readout was drawn on b_0_ DTI images (directly beneath the scalp location where we placed the LIFUS transducer) and masked with both grey-matter and white-matter segments (obtained from steps below) to maintain only the intersection. We used the mean MD in this masked ROI to calculate the percentage ADC change, and recorded occurrences of ADC dropping equal or more than the minimal clinically significant 20%^7^.

The segmentation and co-registration step started with lesion delineations, performed on FLAIR images under the supervision of a stroke neurologist (WF). A lesion was defined as areas of abnormal signal intensities standing out against the surrounding normal tissue, and using our previously described procedures^23^. These lesion masks were paired with T1 images for segmentations by cost-function non-linear registration to an elderly template using the Clinical Toolbox^24^ in Statistical Parametric Mapping 12 (SPM12, University College London, London, UK). The resultant segments were co-registered into the participant’s diffusion space using the normalized mutual information pipeline in SPM12, and these co-registered segments were fed to the ADC calculation above.

#### MSL response time

We quantified motor learning improvements by calculating the percentage change in the within-block median response time. The response time was defined as the time from a cue presentation (circle appearance) to a motor response (button pressing). We calculated the median response time for the five key presses in each sequence repetition (one response time for each repetition) and took the median of these twelve within-repetition medians as the overall median response time for that MSL block. Next, we calculated the percentage post-minus-pre change in the within-block median response time and recorded the number of participants who improved ≥20% on MSL^25^.

#### Corticospinal excitability assessment

In the TMS-MEP, we first identified MEPs as signals that had a biologically plausible latency between the stimulus artifact and the first prominent post-stimulus deflection. Next, we took the peak-to-peak amplitude of each MEP. After sorting these values from low to high, we removed the four outliers from both tails and calculated the mean of the middle twelve (5^th^ – 16^th^) values. The mean amplitude of these twelve MEPs was used as the measure of corticospinal excitability. We then calculated the percentage post-minus-pre change and recorded the number of participants who improved ≥20% on MEP^26^.

#### Statistical Analyses

We summarized participant demographics with descriptive statistics, mean±standard deviation (SDev). To test the effect of intensity on MSL and TMS measures, we lumped 0/1/2 W/cm^2^ participants in one dose group (LOW), and 4/6/8 W/cm^2^ participants in another dose group (HIGH). Metrics of comparison, which were presented median±SDev, were a) the percentage improvement in MSL response time and b) the percentage variation in the peak-to-peak amplitude of MEPs. Given the sample size of eighteen participants, we performed the non-parametric Fligner-Policello robust rank order test with bootstrapping to not assume data normality or symmetry^27^. For categorical analysis, a Fisher’s Exact test was used to compare the two groups. Tests were performed in MATLAB (R2020a, MathWorks, Inc., MA, USA), significance level = 0.05.

## RESULTS

Thirty stroke participants were screened between October 2021 and May 2024. Eighteen participants (6 females, aged 52±14 years) met the eligibility criteria and participated in the study **(Table 1A)**. Each participant received one stimulation dose/intensity, with three participants at each dose as the intensity was escalated to 8 W/cm^2^ without meeting pre-defined stopping rules. No participant had a ≥2^nd^-degree scalp burn or clinical seizure, visible lesion on the DTI, clinically significant (≥ 20% reduction) ADC reduction **(Figure 3)**, and discontinuation **(Table 1B)**. Since no major response occurred at any intensity, the trial was completed with eighteen participants. Although no stopping rules were met, one participant (4 W/cm^2^) suffered a first-degree scalp burn with mild pain sensation that resolved on the next day.

**Figure 3.**
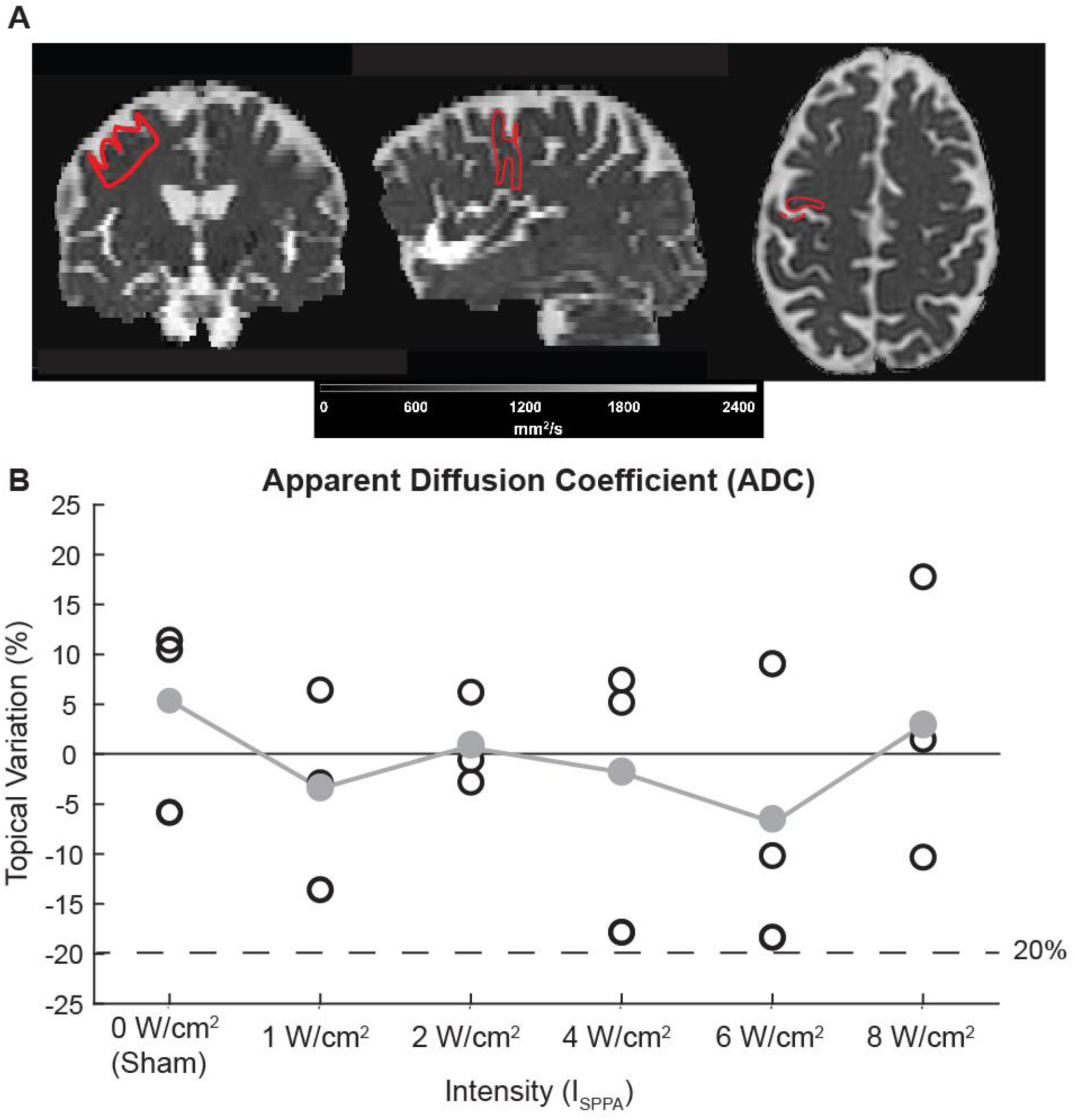
Topical apparent diffusion coefficient variations. **A**, Sample mean diffusivity map (Participant 11) overlaid with region of interest outline (red). **B**, Individual (white circle) and within-intensity mean (gray circles connected with line) variation of apparent diffusion coefficient against intensity. μm^2^/ms: square micrometers per millisecond; W/cm^2^: Watts per square centimeter.

**Table 1.** A, Baseline participant characteristics. B, Safety outcomes. A.

| Participant ID | Intensity, W/cm<sup>2</sup> | Sex, F/M | Race, A/B/O/W | Stroke type, I/H | Stroke hemisphere, L/R | Time from stroke, m | NIH Stroke scale | FM-UE scale | Affected side RMT, % MSO | Unaffected side RMT, %MSO |
| --- | --- | --- | --- | --- | --- | --- | --- | --- | --- | --- |
| 1 | 0 | F | B | H | R | 39 | 2 | 59 | 56 | 61 |
| 2 |  | M | B | I | L | 11 | 1 | 60 | 71 | 49 |
| 3 |  | M | B | I | L | 27 | 0 | 57 | 45 | 38 |
| 4 | 1 | M | B | H | R | 87 | 1 | 50 | 59 | 54 |
| 5 |  | M | A | H | R | 3 | 0 | 58 | 50 | 36 |
| 6 |  | F | B | I | R | 5 | 0 | 58 | 45 | 40 |
| 7 | 2 | F | W | I | R | 25 | 0 | 60 | 68 | 55 |
| 8 |  | M | W | I | L | 120 | 0 | 61 | 50 | 54 |
| 9 |  | M | B | I | L | 6 | 5 | 51 | 48 | 49 |
| 10 | 4 | F | W | I | R | 26 | 2 | 55 | 70 | 37 |
| 11 |  | M | W | I | R | 12 | 2 | 61 | 40 | 37 |
| 12 |  | M | W | H | R | 29 | 4 | 38 | 54 | 44 |
| 13 | 6 | F | W | I | R | 37 | 1 | 60 | 53 | 60 |
| 14 |  | M | W | I | R | 8 | 1 | 62 | 57 | 66 |
| 15 |  | F | B | I | L | 5 | 4 | 55 | 55 | 57 |
| 16 | 8 | M | W | I | R | 12 | 3 | 56 | 37 | 41 |
| 17 |  | M | W | H | R | 1 | 2 | 61 | 42 | 40 |
| 18 |  | M | B | I | R | 6 | 0 | 61 | 57 | 49 |

**B**
| Participant ID | Intensity, W/cm<sup>2</sup> | 2<sup>nd</sup>-degree scalp burn, Y/N | Seizure, Y/N | ADC reduction ≥25%, Y/N | Discontinuation due to any reason, Y/N |
| --- | --- | --- | --- | --- | --- |
| 1 | 0 | N | N | N | N |
| 2 |  | N | N | N | N |
| 3 |  | N | N | N | N |
| 4 | 1 | N | N | N | N |
| 5 |  | N | N | N | N |
| 6 |  | N | N | N | N |
| 7 | 2 | N | N | N | N |
| 8 |  | N | N | N | N |
| 9 |  | N | N | N | N |
| **10** | **4** | **N** | **N** | **N** | **N** |
| **11** |  | **N** | **N** | **N** | **N** |
| **12** |  | **N** | **N** | **N** | **N** |
| **13** | **6** | **N** | **N** | **N** | **N** |
| **14** |  | **N** | **N** | **N** | **N** |
| **15** |  | **N** | **N** | **N** | **N** |
| **16** | **8** | **N** | **N** | **N** | **N** |
| **17** |  | **N** | **N** | **N** | **N** |
| **18** |  | **N** | **N** | **N** | **N** |

W/cm<sup>2</sup>: Watts per square centimeter; F: female; M: male; Yr: years; A: Asian; B: Black; O: Others; W: White; I: ischemic; H: hemorrhagic; L: left; R: right; m: months; NIH: National Institutes of Health; FM-UE: Fugl-Meyer upper extremity; RMT: resting motor threshold; %MSO: percentage maximum stimulator output; Y: yes; N: no; ADC: apparent diffusion coefficient. HIGH intensity (bold font).

### MSL

Sixty-seven percent of participants (6/9) in HIGH improved ≥20% on the MSL, whereas no participant (0/9) in LOW did, *p*=0.009. Similarly, the median percentage MSL improvement of HIGH (24.7±13.3%) was statistically significantly higher than that in LOW (13.2±10.9%), *p*=0.014 **(Figure 4A)**.

**Figure 4.**
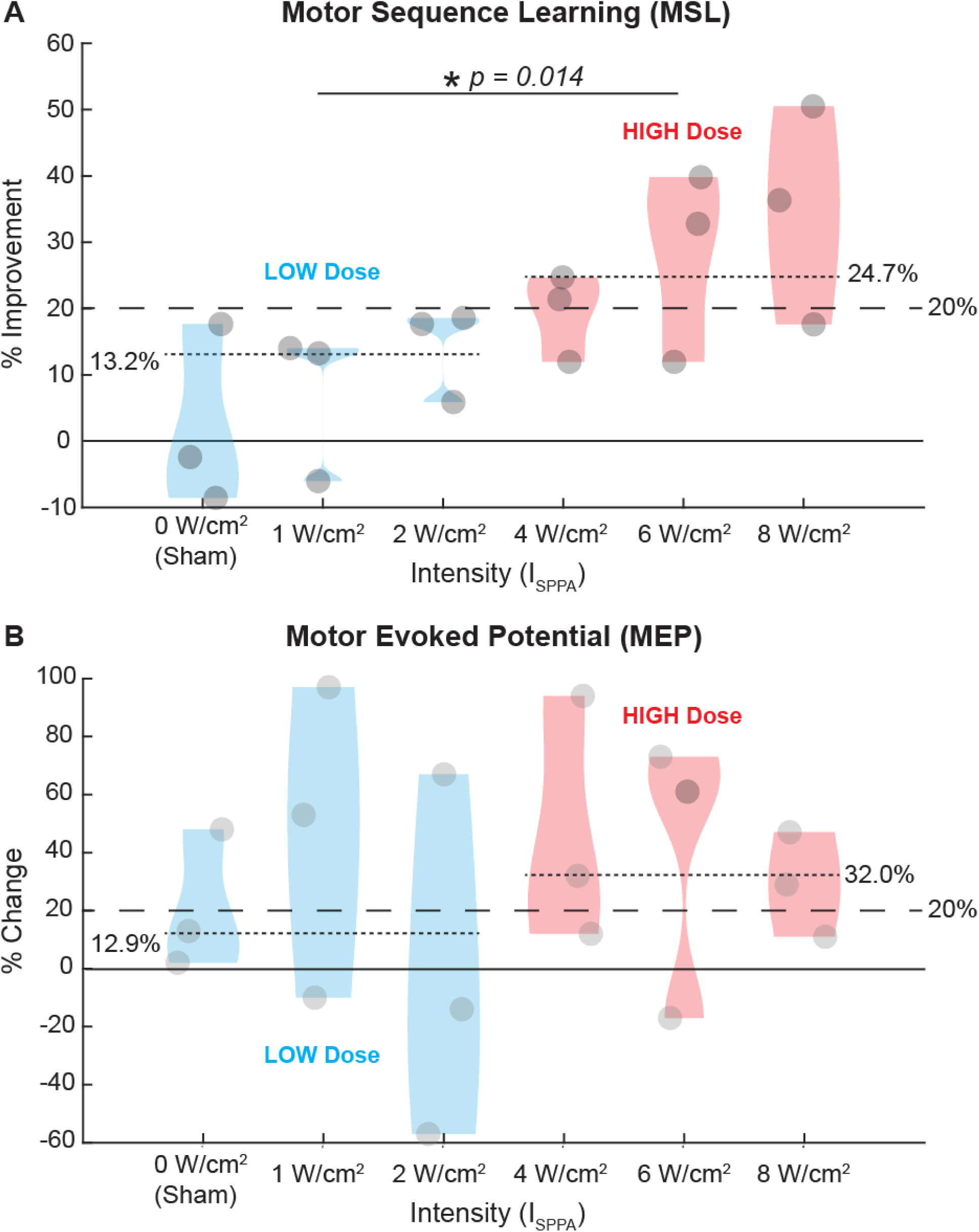
Behavioral and neurophysiological outcomes. **A**, Percentage MSL improvements against intensities. Positive indicates faster response time. 67% (6/9) of participants in HIGH improved ≥ 20% on the MSL, whereas no participant (0/9) in LOW did (*p*=0.009). Similarly, percentage MSL improvements in LOW are significantly different from the HIGH, *p*=0.014. **B**, Percentage MEP changes against intensities. Positive indicates greater corticospinal excitability. 67% (6/9) in HIGH had ≥ 20% enhanced corticospinal excitability as compared to 44% (4/9) in LOW (*p*=0.67). Similarly, percentage MEP changes in LOW are not significantly different from HIGH, *p* = 0.53. **A-B**, Horizontal dashed lines identify a 20% change in MSL/MEP amplitude, both the likely minimal clinically meaningful change^25,26^, horizontal dotted lines indicate group medians for HIGH and LOW, and circles are individual data points. W/cm^2^: Watts per square centimeter; SDev: standard deviation.

### TMS

Sixty-seven percent of participants (6/9) in HIGH, compared to forty-four percent (4/9) in LOW, had enhanced corticospinal excitability ≥20%, *p*=0.67. The median percentage increase in MEP amplitude for HIGH (32.0±34.3%) was not statistically significantly higher than that in LOW (12.9±48.0%), *p*=0.53 **(Figure 4B)**.

## DISCUSSION

To the best of our knowledge, this study is the first to test the potential application of LIFUS in stroke patients for motor implications. Our data is critical and instrumental for the stroke recovery field to further establish the efficacy of LIFUS in stroke population. There were zero occurrences of pre-defined major adverse events. All participants tolerated LIFUS well and completed all study procedures. We used ADC qualitatively and quantitatively to ensure safety at subclinical safety level. Qualitatively, there is no participant with visible lesion on the DTI sequence. Quantitively, there is no participant with ADC reduction ≥ 20% or absolute ADC threshold^28^ ≤620×10^−6^ mm^2^/s. One participant had a mild first-degree scalp burn, but symptoms went away the next day without seeking medical attentions. An insufficient amount of ultrasound gel was likely the cause. Simulation data suggests that our intensity/dose range should have minimal scalp temperature rises. However, this does highlight the need to further monitor safety issues in future studies with multiple sessions.

We used a traditional 3+3 dose-escalation trial design (the modified Fibonacci method) and demonstrated that single-session, 12-minute LIFUS up to 8 W/cm^2^ I_SPPA_ is safe and tolerable in stroke participants. The outcome of 3+3 trials is that the maximum tolerable dose (intensity) is the intensity/dose at which 1/3 or more of participants experience pre-defined dose (intensity)-limiting major response(s) (i.e., meeting the stopping dose). This design is simple, clear, and has been accepted and widely used in drug trials, although the selection of one-third or more for stopping rule is admittedly arbitrary. The alternative is the accelerated titration design^29^ which starts at a low dose, assesses its toxicity degree, and uses that extent to determine the next dose. This accelerated titration design is suitable for wide dose range and no initial approximate endpoint. Adhering the FDA and the International Electrotechnical Commission guidelines^15,30^, the maximum I_SPPA_ for LIFUS is around 8 W/cm^2^. Therefore, the modified Fibonacci method was deemed appropriate. It remains unknown whether we should escalate beyond the current intensity limit, which is largely for diagnostic ultrasounds. Escalating beyond FDA guidelines surely needs to be done under an investigational device exemption. While no participant experienced any pre-defined major responses, there is nevertheless a small possibility that it was due to a statistical chance. We do believe our trial conceptually establishes the safety and tolerability of LIFUS in stroke patients.

In addition to determining the safety of LIFUS in stroke patients, we also observed effects of LIFUS on motor learning and corticospinal excitability in the stroke population. LIFUS at ipsilesional M1 during a motor practice can enhance motor skill learning, with more significant effects at higher intensities – 67% (6/9) participants in HIGH had ≥ 20% improvement in MSL while no participant in LOW had so. Corticospinal excitability showed the same direction as the motor learning effect, but the magnitude is relatively small and does not reach significance. There are several potential explanations. Our study is not powered to detect a significance in corticospinal excitability variations. MEPs are known to have a huge variability in both healthy controls^31^ and in stroke patients with an injured corticospinal tract^32^, even though we incorporated several mitigating approaches. For example, we waited over ten seconds between TMS pulses, which is known to reduce MEP variabilities^20^. The huge individual variability requires a large sample size to detect a significant change in MEPs. It was reported that, to reliably detect a 20% difference in MEP amplitudes, each group requires thirty participants^26^. Nevertheless, what really matters is that corticospinal excitability moved in the same direction as the motor learning effect.

Our result is consistent with but also different from several studies investigating LIFUS and its influence on the corticospinal excitability^33^. Two prior reports were conducted with healthy volunteers^5,33–37^ and one with Parkinson’s disease participants^34^, all with the brain intact. Our study population had structural brain lesion(s) with corresponding motor deficits. Also, parameters other than the intensity, such as PRF, duty cycle and SD, could be important factors too. Samuel et al. (2023) used an average I_SPTA_ about 10% of ours, but found it induces cortical excitability in Parkinson’s disease participants^34^. This group used a much lower PRF (5 Hz vs. 1000 Hz) and a much longer SD (80 s vs. 500 ms). In another example, Zhang et al. (2023) showed that LIFUS with different PRFs, but both 2.46 W/cm^2^ I_SPPA_, can induce differential cortical excitability in healthy controls^38^, whereas our study at a much higher I_SPPA_ (8 W/cm^2^) with a 1000 Hz PRF enhanced corticospinal excitability in stroke patients. We did not observe a trend of reduced corticospinal excitability in LOW. **Table 2** summarizes ultrasound parameters from various studies and their effect on the corticospinal excitability. It further underscores the necessity and importance of parameter optimization and individualization in specific disease populations.

**Table 2.**
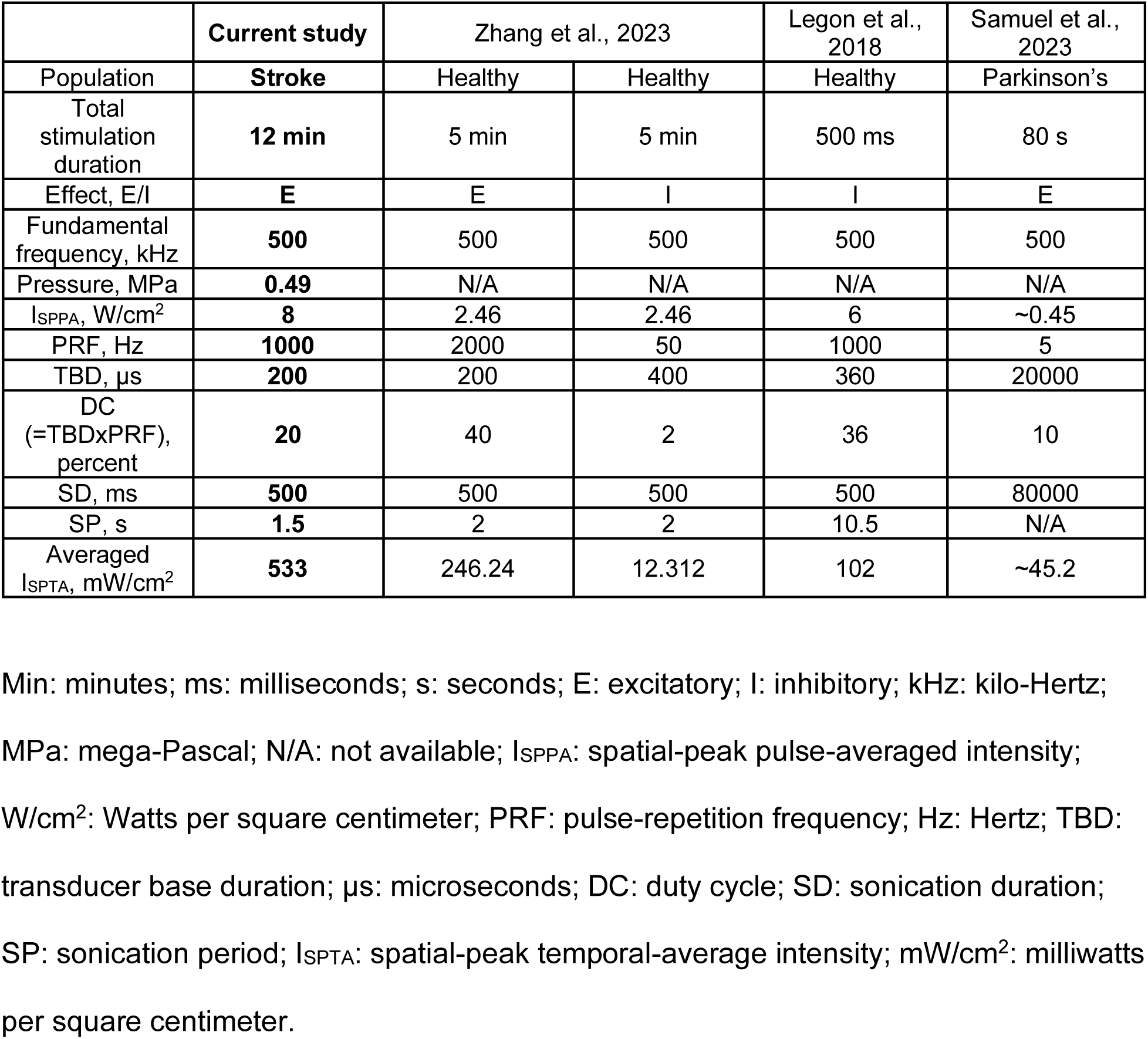
Ultrasound parameters in different studies.

Our study is not free from limitations. First, although we demonstrated the safety and tolerability of single 12-minute, 500 kHz and up to 8 W/cm^2^ I_SPPA_ LIFUS using a 3+3 trial, cautions should be exercised when interpreting the data. Serious adverse events may occur at very low odds. Zero major responses could simply be due to a statistical chance with eighteen participants. One statistical simulation demonstrates that about 30% of 3+3 trials identify the maximum tolerated dose^39^. Safety profile should continue to be monitored in future studies in order to protect vulnerable stroke patients. Secondly, we only tested intensity (I_SPPA_) as the major LIFUS parameter in this safety study and controlled other parameters. Parameters such as PRF, DC and SD may also influence learning behaviors and corticospinal excitabilities, prompting a need for further investigations. Establishing optimal parameters is fundamental and critical for LIFUS to successfully treat certain neuropsychiatric diseases.

## CONCLUSIONS

In this Phase I study, we demonstrated that a 12-minute session of 500 kHz LIFUS up to 8 W/cm^2^ I_SPPA_ is feasible, safe, and tolerable in stroke participants. Also, stroke participants receiving higher intensity LIFUS (HIGH) performed significantly better on the MSL task and showed signs of greater LIFUS-induced corticospinal excitability, compared to participants receiving lower intensity LIFUS (LOW). A logical next step is to further assess the safety and preliminary efficacy of LIFUS in a Phase II study to systematically investigate LIFUS in stroke patients with motor or other impairments

## Data Availability

The present study is supported by the American Heart Association (AHA) and will adhere to the data sharing policies of AHA.

## Acknowledgments

Authors thank all stroke participants who volunteered.

